# Exact results for the stochastic SIS epidemic model in densely populated environments

**DOI:** 10.1101/2025.08.11.25333426

**Authors:** Tingting Chen, Zhen Jin, Guirong Liu, Chen Jia

## Abstract

In this study, we investigate the stochastic dynamics of an extended SIS epidemic model in densely populated environments within a Markov jump process framework. We solve the master equation in closed form and obtain exact solutions of the time-dependent distribution of the number of infected individuals, the quasi-stationary distribution, the extinction time distribution of the epidemic, and the distribution of the first-passage time at which the number of infections reaches a certain threshold. The approximated quasi-stationary distribution and mean extinction time are also derived using the large deviation theory. Interestingly, we find that the first nonzero eigenvalue of the generator matrix of the Markovian model characterizes the extinction rate of the epidemic, while the second nonzero eigenvalue characterizes its outbreak rate. We also examine the stochastic bifurcation for our model based on the time evolution of the probability distribution and the bifurcation threshold of the basic reproduction number for the stochastic SIS model is shown to be large than that for its deterministic counterpart. Finally, we demonstrate that analyzing the first-passage time distribution can offer early warning for interventions and optimize the allocation of emergency beds.

## 1 Introduction

The spread of infectious diseases poses a significant threat to human populations, making it a critical concern for public health [1, 2]. Epidemic models have become essential tools for understanding the transmission mechanisms of infectious diseases, providing a theoretical framework to analyze past outbreaks and predict the future trajectory of epidemics. Common epidemic models include the SI (susceptible-infected), SIS (susceptible-infected-susceptible), and SIR (susceptible-infected-removed) compartment models, which typically classify individuals into different compartments based on their disease status [3–5]. These models are chosen based on the pathological characteristics of specific epidemics.

Over the past three decades, significant progress has been made in the mathematical modeling and theoretical analysis of various epidemic models, extensively studied from both deterministic and stochastic perspectives. Classical epidemic models assume homogeneous mixing within the population and disease transmission occurs through effective contact between susceptible and infected compartments. When the population size is very large, random fluctuations in disease transmission can be ignored, and the evolution of the number of individuals in each compartment is typically modeled by a set of ordinary differential equations (ODEs). Research within this framework has focused on various aspects such as threshold dynamics [6], bifurcation analysis [7], parameter inference [8], and optimal control [9]. The classical framework has also been enriched through the integration of network topologies [10], spatial diffusion [11, 12], time delays [13], and intervention measures [14], enabling deeper insights into the spread of epidemics.

However, when the population size is relatively small, random fluctuations in disease transmission can no longer be ignored, and the evolution of the system should be modeled within a stochastic framework. There are typically two types of stochastic epidemic models: stochastic differential equation (SDE) models and continuous-time Markov chain (CTMC) models [5, 15, 16]. In SDE models, the population size in each compartment is treated as a real-valued continuous variable, whereas in CTMC models, the population size is treated as an integer-valued discrete variable, and hence provides a more accurate description. The probability distribution of the population size in each compartment is governed by a Fokker-Planck equation for SDE models and by a master equation for CTMC models. At the center of the stochastic dynamic theory of infectious diseases is a limit theorem proved by Kurtz in the 1970s [17, 18], which states that, in the limit of large population size, the CTMC model converges to the deterministic ODE model with probability one over any finite time interval. Roughly speaking, in Kurtz’s theory, ODE models can be viewed as first-order approximations of CTMC models when the population size is large, while SDE models serve as second-order approximations (also referred to as diffusion approximations) of CTMC models [18].

Due to the complexity of CTMC models, various approximation techniques have been developed, including diffusion approximations [18, 19], van Kampen’s system-size expansion [20], Wentzel-Kramers-Brillouin (WKB) approximations [21, 22], and branching process approximations [23, 24]. In general, the first three approximation methods provide accurate predictions for relatively large population sizes, whereas branching process approximations are mainly valid during the initial stages of an epidemic. These techniques have been extensively used to study key topics such as threshold dynamics [15, 16], stochastic bifurcation analysis [25, 26], moment closure strategies [27], estimation of the time to extinction [28, 29], and quasi-stationary distribution prior to disease extinction [30, 31].

While various approximation techniques have been extensively studied, exact results for stochastic epidemic models remain quite limited due to the nonlinear interactions between compartments. In this work, we present a systematic study of an extended stochastic SIS model in crowded environments within a CTMC framework. We derive closed-form solutions for several important quantities, including the time-dependent distribution of the number of infected individuals, the quasi-stationary distribution, the extinction time distribution of the epidemic, and the first-passage time distribution. The classical SIS model describes the dynamics of infectious diseases where recovered individuals do not develop immunity and can be re-infected after recovery. However, in densely populated areas, such as public transportation, schools, and hospitals, an infected individual may simultaneously infect multiple susceptible individuals. Such non-pairwise interaction mechanisms in disease transmission are sometimes modeled and analyzed using simplicial contagion models [32–34]. Our work extends the classical SIS model by incorporating simultaneous transmission to multiple individuals, making it more suitable for infectious diseases that spread rapidly in densely populated and crowded environments. This generalization significantly increases the mathematical complexity, since the spectra of the classical SIS model are all real, whereas those of the extended SIS model may be complex.

The paper is organized as follows. In Sec. 2, we propose an extended stochastic SIS model in densely populated environments and compare it with its deterministic counterpart. In Sec. 3, we analytically solve the master equation of the stochastic model and derive the exact time-dependent distribution of the number of infected individuals expressed in terms of the eigenvalues of the generator matrix. We find that the time evolution of the probability distribution may exhibit two distinct patterns, and the stochastic bifurcation between these patterns is examined in detail. In Sec. 4, we use the analytical time-dependent distribution to obtain the exact quasi-stationary distribution, which is then analyzed and compared with the WKB approximation. In Sec. 5, we investigate the extinction rate and extinction time of the epidemic. We find that the first nonzero eigenvalue of the generator matrix characterizes the extinction rate of the epidemic, while the second nonzero eigenvalue characterizes its outbreak rate. Additionally, we also derive the exact distribution and expected value of the extinction time and compare them with the WKB approximation. In Sec. 6, we derive the exact distribution of the first-passage time at which the number of infections reaches a given threshold, and we demonstrate how analyzing this distribution can guide infectious disease control. We conclude in Sec. 7.

## 2 An extended SIS model

### 2.1 Classical SIS model

In some infectious diseases, such as gonorrhea, chlamydia, influenza, the common cold, and respiratory syncytial virus, infection does not confer long-lasting immunity [35–37]. Individuals who recover from the disease return to the susceptible state immediately and can be infected again. Such diseases are typically modeled within the classical SIS framework (Fig. 1(a)). Specifically, we consider a closed population of size *N*, where each individual is classified as either susceptible or infected. Let *N*_*S*_(*t*) and *N*_*I*_ (*t*) denote the numbers of susceptible and infected individuals at time *t*, respectively, such that *N*_*S*_(*t*) + *N*_*I*_ (*t*) = *N*. The densities of the two subpopulations (compartments) are then given by

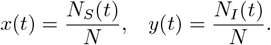

**Figure 1:**
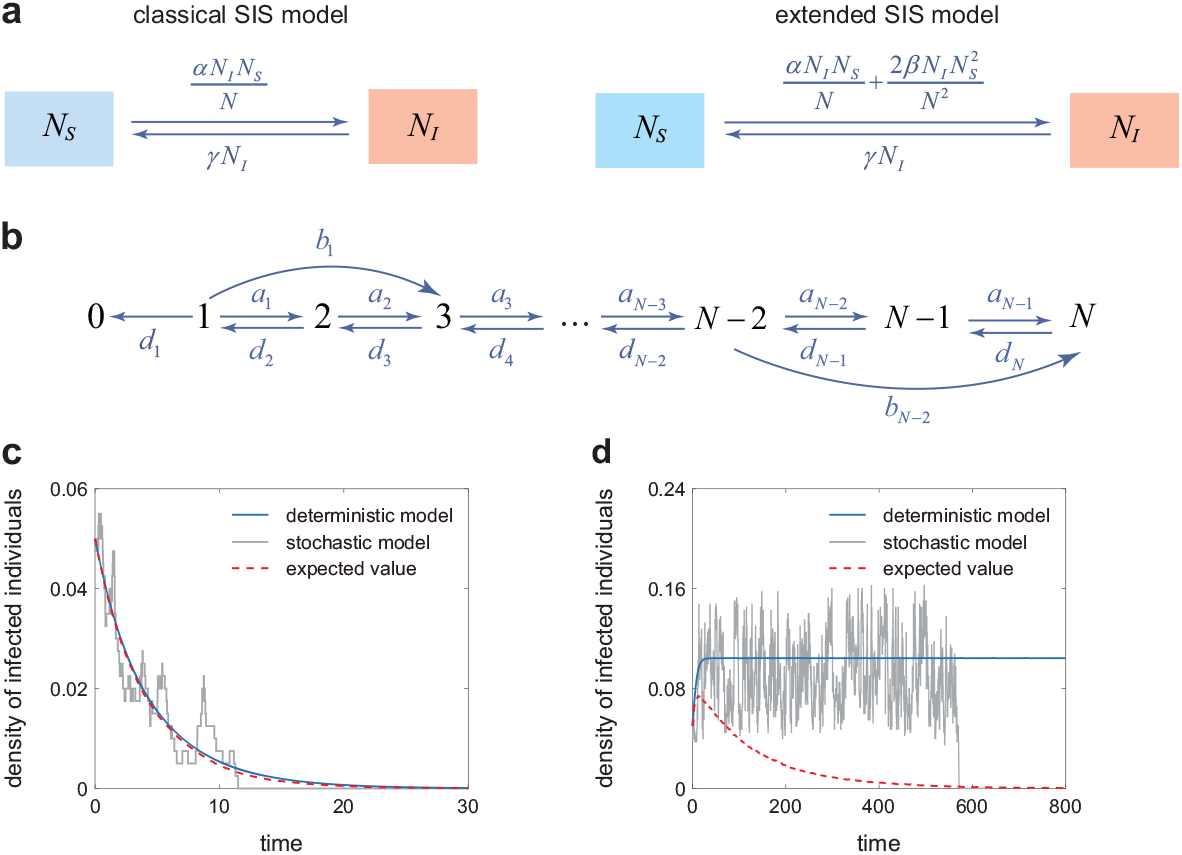
Deterministic and stochastic SIS epidemic models. **(a)** Illustration of the classical and extended deterministic SIS models. **(b)** Transition diagram of the Markovian dynamics for the stochastic (extended) SIS model. **(c)** Comparison of the trajectory of *x*(*t*) for the deterministic SIS model (blue curve), the trajectory of *N*_*I*_ (*t*)*/N* for the stochastic SIS model (grey curve), and the expected value of *N*_*I*_ (*t*)*/N* for the stochastic SIS model (red curve) when *R*_0_ *<* 1. The model parameters are chosen as *α* = 0.4, *β* = 0.2, *γ* = 1, *R*_0_ = 0.8, and *N* = 400. **(d)** Same as in (c) but for *R*_0_ *>* 1. The model parameters are chosen as *α* = 0.4, *β* = 0.4, *γ* = 1, *R*_0_ = 1.2, and *N* = 400. In (c),(d), the deterministic trajectory is the solution to Eq. (2), the stochastic trajectory is generated using Gillespie’s stochastic simulation algorithm, the expected value is computed as the sample mean of 10^4^ stochastic trajectories, and the initial condition is chosen as *N*_*I*_ (0) = 0.05*N*.

To proceed, let *α* be the disease transmission rate, i.e. the average number of contacts per person per unit time that result in disease transmission between an infected and a susceptible individual. Since an infected individual contacts a susceptible individual with probability *N*_*S*_*/N*, the average number of new infections per unit time is *αN*_*I*_*N*_*S*_*/N*. In addition, let *γ* denote the recovery rate of an infected individual. Then *γN*_*I*_ represents the average number of infected individuals recovering from the disease per unit time. It was recently pointed out [38] that the above epidemic model is essentially equivalent to a chemical reaction system governed by mass-action kinetics, where random collisions between molecules result in reactions, and the reaction rate is proportional to the concentrations of the reactants. Specifically, the classical SIS model corresponds to the following chemical reaction system [39]:

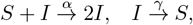

where *S* denotes a susceptible individual and *I* denotes an infected individual. The first reaction represents the infection of a susceptible individual, while the second reaction represents the recovery of an infected individual.

When the total population size *N* is very large, the evolution of the numbers of susceptible and infected individuals is described by the deterministic SIS model

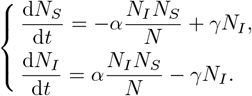

Since the total population size *N*_*s*_ + *N*_*I*_ = *N* remains constant, the above system can be reduced to the following single-variable ODE:

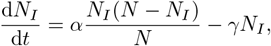

which describes the evolution of the number of infected individuals. Sometimes, it is more convenient to consider the density of infected individuals, *x*(*t*) = *N*_*I*_ (*t*)*/N*, whose evolution is governed by the ODE

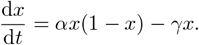

### 2.2 Extended SIS model

The classical SIS model assumes that each infected individual can transmit the disease to only one susceptible individual per contact. However, in densely populated settings, such as public transportation, schools, and hospitals, the number of individuals involved in each contact can increase significantly. This may result in multiple individuals being infected simultaneously during a single contact event. To capture this phenomenon, we propose an extended SIS model in which each infected individual can simultaneously contact two susceptible individuals and transmit the disease to both (Fig. 1(a)). Specifically, let *β* denote the rate of simultaneous infection, i.e. the average number of contacts per person per unit time that result in simultaneous transmission of the disease to two susceptible individuals. It is clear that an infected individual comes into contact with two susceptible individuals with probability

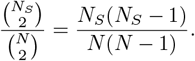

Hence the average number of new infections resulting from simultaneous transmission per unit time is given by

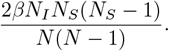

Similarly, the extended SIS model is equivalent to the chemical reaction system

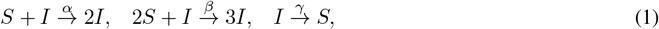

where the first reaction represents disease transmission to a single susceptible individual, the second reaction describes simultaneous transmission to two susceptible individuals in crowded environments, and the third corresponds to the recovery of an infected individual. Note that the extended SIS model reduces to the classical one when *β* = 0. Similar reaction system has also be used to model the regulation of stem cell dynamics [40].

When the total population size *N* is very large, the evolution of the numbers of susceptible and infected individuals is described by the following system of deterministic ODEs:

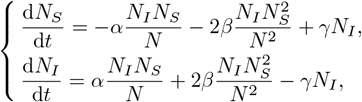

where we have used the approximations of 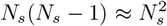 and *N* (*N* − 1) ≈ *N* ^2^ when *N* ≫ 1. Since the total population size *N*_*s*_ + *N*_*I*_ = *N* is conserved, the evolution of the density of infected individuals, *x*(*t*) = *N*_*I*_ (*t*)*/N*, is governed by

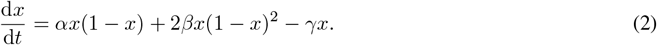

Clearly, the interval [0, 1] is positively invariant with respect to this system, and the system must have a disease-free equilibrium *x* = 0. Using the next generation matrix approach [41], it is easy to see that the basic reproduction number of the extended SIS model is given by

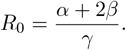

When *R*_0_ ≤ 1, the disease-free equilibrium globally asymptotically stable within [0, 1]; when *R*_0_ *>* 1, the disease-free equilibrium is unstable and the system also has a positive endemic equilibrium

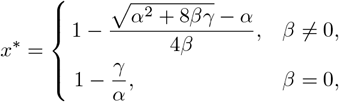

which is globally asymptotically stable within (0, 1]. In what follows, unless otherwise stated, we always focus on the extended SIS model since it provides a more accurate description of how infectious diseases spread in densely populated environments than the classical SIS model.

### 2.3 Stochastic SIS model

When the total population size *N* is relatively small, stochasticity cannot be ignored and it is necessary to consider a stochastic epidemic model where the numbers of susceptible and infected individuals take integer values rather than real values as in the deterministic model. When stochastically modeled, the evolution of the extended SIS system is governed by a Markov jump process (*N*_*S*_(*t*), *N*_*I*_ (*t*))_*t*≥0_ with state space {(*m, n*) ∈ ℕ^2^|*m* + *n* = *N* }, where *m* and *n* denote the numbers of susceptible and infected individuals, respectively. Clearly, the Markovian model can only make the following three types of transitions:

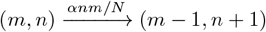

due to disease transmission to a single susceptible individual,

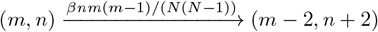

due to simultaneous transmission to two susceptible individuals, and

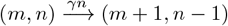

due to recovery of an infected individual. Note that here the transition rates for the three events are exactly the same as those in the deterministic model (Fig. 1(a)). Since *N*_*S*_(*t*) + *N*_*I*_ (*t*) = *N*, we only need to consider the dynamics of the single-variable process *N*_*I*_ (*t*), which is a Markov jump process with state space *S* = {0, 1, …, *N* }. Similarly, the process *N*_*I*_ (*t*) can make the following three types of transitions:

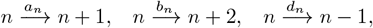

with transition rates

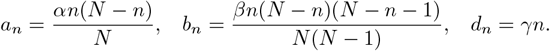

The transition diagram of the process *N*_*I*_ (*t*) is shown in Fig. 1(b). Clearly, we have *a*_0_ = *b*_0_ = *d*_0_ = 0 and thus 0 ∈ *S* is an absorbing state. The other states 1, …, *N* are all non-absorbing and form a communicating class [42]. Let *P*_*n*_(*t*) denote the probability of having *n* infected individuals at time *t*. Then the evolution of the Markovian model is governed by the Kolmogorov forward equation (also known as the master equation)

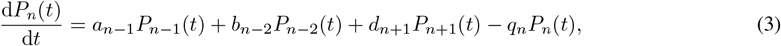

where *q*_*n*_ = *a*_*n*_ + *b*_*n*_ + *d*_*n*_ denotes the total transition rate from state *n* to all other states and *P*_*n*_(*t*) ≡ 0 by default for any *n* ∉ *S*. Let *P* (*t*) = (*P*_0_(*t*), …, *P*_*N*_ (*t*)) denote the probability distribution of the number of infected individuals at time *t*. Then the master equation can be written in matrix form as

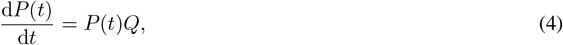

where

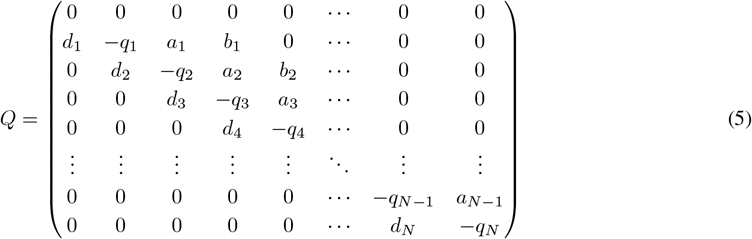

is the generator matrix of the Markovian model. Since 0 ∈ *S* is an absorbing state, the first row of *Q* is the zero vector.

The relationship between the deterministic ODE model and the stochastic CTMC model was rigorously proved by Kurtz [17, 18]. Let *N*_*I*_ (*t*) denote the number of infected individuals for the stochastic model and *x*(*t*) denote the density of infected individuals for the deterministic model. Kurtz’s theorem states that for any *T >* 0,

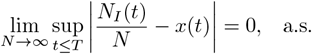

The theorem states that, in the limit of large population size, the density of the stochastic model converges to that of the deterministic model over any finite time interval with probability one. We emphasize that although the deterministic and stochastic models exhibit similar dynamic behavior over finite time periods, their long-term behavior can be fundamentally different. Since the Markov jump process *N*_*I*_ (*t*) has a finite state space with 0 ∈ *S* being the unique absorbing state, the ergodic theorem implies that the disease will eventually go extinct with probability one for the stochastic SIS model. This is in sharp contrast to the predictions of the deterministic model, in which the density of infected individuals converges to a positive endemic equilibrium *x*^∗^ when *R*_0_ *>* 1.

Fig. 1(c),(d) compare the time evolution of the density of infected individuals *x*(*t*) for the deterministic SIS model, the time evolution of the density of infected individuals *y*(*t*) = *N*_*I*_ (*t*)*/N* for the stochastic SIS model, and the time evolution of the expected value of *y*(*t*) for the stochastic SIS model. Clearly, the two models have similar dynamic behaviors when *R*_0_ *<* 1. However, the long-term behaviors for the two models are totally different when *R*_0_ *>* 1 — the deterministic trajectory tends to the positive endemic equilibrium *x*^∗^, while the stochastic trajectory first fluctuates around the deterministic trajectory over a finite period of time, and eventually becomes zero due to extinction of the disease. Moreover, the expected value *E* [*y*(*t*)] of the density of infected individuals tends to zero for the stochastic model, which is inconsistent with the predictions of the deterministic model (Fig. 1(d)).

## 3 Time-dependent distribution

### 3.1 Exact solution

We next focus on the time evolution of the stochastic SIS model by computing the probability distribution of the number of infected individuals at each time. Understanding the time evolution of the distribution provides insight into how the disease spreads. Following the method proposed in [43], we next solve the master equation in time and thus obtain the time-dependent distribution *P*_*n*_(*t*) of infected individuals. The solution to Eq. (4) can be represented using matrix exponential as

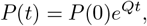

where *P* (0) = (*P*_0_(0), …, *P*_*N*_ (0)) denotes the initial distribution of infected individuals. In general, it is difficult to compute the matrix exponential exactly since we need to calculate the eigenvalues and eigenvectors of the generator matrix *Q*. However, the special structure of *Q* allows us to bypass the eigenvector calculation. By Cauchy’s integral formula for matrices [44], for any continuous function *f*, we have

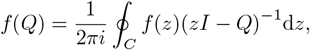

where *C* is an arbitrary simple closed curve in the complex plane that contains all the eigenvalues of *Q* in its interior. If we take *f* (*z*) = *P* (0)*e*^*zt*^, then we obtain

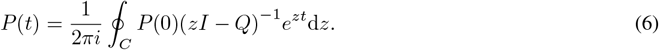

Suppose that the initial number of infected individuals is *n*_0_ ∈ *S*; then the initial distribution is given by 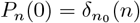, where 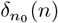 a Kronecker delta that takes the value of 1 when *n* = *n*_0_ and the value of 0 otherwise. We will explain later how to extend our results to more general initial distributions. Under the point initial distribution, Eq. (6) can be simplified as

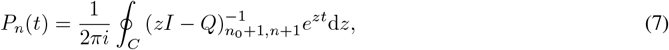

where 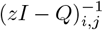 denotes the element of the matrix (*zI* − *Q*)^−1^ in the *i*th row and the *j*th column. By Cramer’s rule of calculating the inverse matrix, we can prove that

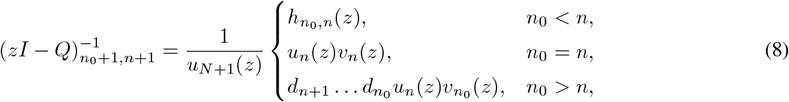

where *u*_*n*_(*z*), 0 ≤ *n* ≤ *N* + 1, are polynomials defined recursively by

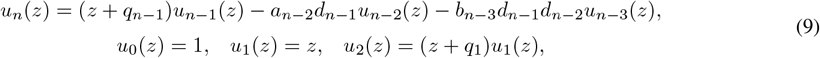

with *u*_*N*+1_ = det(*zI* − *Q*) being the characteristic polynomial of *Q, v*_*n*_(*z*), 0 ≤ *n* ≤ *N*, are polynomials defined recursively by

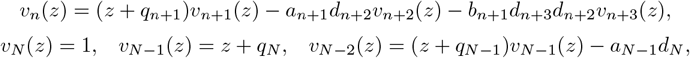

and 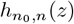 are functions defined as

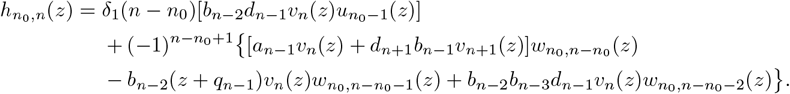

Here 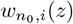, 1 ≤ *i* ≤ *n* − *n*_0_, are another sequence of polynomials defined recursively by

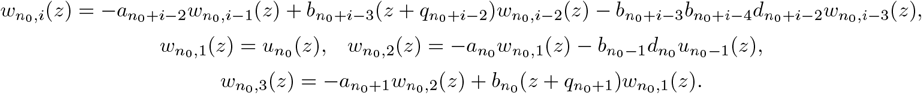

Combining Eqs. (7) and (8), we obtain

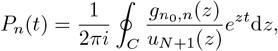

where 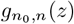 are polynomials defined as

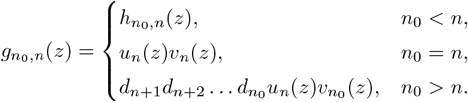

Since *u*_*N*+1_(*z*) is the characteristic polynomial of *Q*, we have

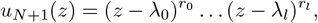

where λ_0_, …, λ_*l*_ are all pairwise distinct eigenvalues of *Q* with *r*_0_, …, *r*_*l*_ being their multiplicities, respectively. We can then apply Cauchy’s residue theorem

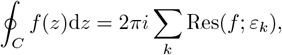

where *ε*_*k*_ are all isolated singularities of *f* inside the simple closed curve *C*. In our current case, all the isolated singularities are the eigenvalues λ_*k*_ and thus

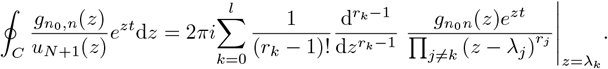

Therefore, once we have known all the eigenvalues of *Q*, the time-dependent solution of Eq. (3), i.e. the time-dependent distribution of infected individuals, is given by

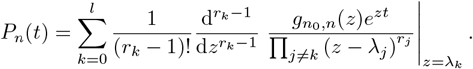

In most cases, the eigenvalues of *Q* are mutually different (in fact, any matrix can be approximated by matrices with non-degenerate eigenvalues to any degree of accuracy). In this case, we have *l* = *N* and *r*_0_ = *· · ·* = *r*_*l*_ = 1, and thus the time-dependent distribution can be simplified as

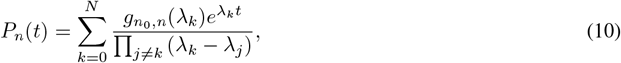

where λ_0_, …, λ_*N*_ are all the eigenvalues of *Q*, i.e. all the zeros of the characteristic polynomial det(*zI* − *Q*). Thus far, we have obtained the exact time-dependent solution of the master equation for the stochastic SIS model under a point initial distribution. In the general case that the system starts from a general initial distribution *p*_*n*_(0) = *π*_*n*_, time-dependent distribution is given by

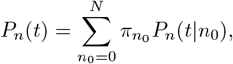

where *P*_*n*_(*t*|*n*_0_) is the time-dependent distribution when the system starts from the fixed state *n*_0_.

As a check of our exact time-dependent solution, we compare it with the numerical one obtained by solving the master equation Eq. (4) directly using the MATLAB function “ODE45” (Fig. 2). As expected, the two solutions agree perfectly at all times. Interestingly, we observe that the stochastic SIS model can produce two different patterns of time evolution. When *R*_0_ is small, the distribution of infected individuals is unimodal at all times (Fig. 2(a)); when *R*_0_ is large, the distribution is unimodal at small and large times, and becomes bimodal with both a zero and a nonzero peak at intermediate times (Fig. 2(b)). The two modes of the bimodal distribution correspond to the disease-free and endemic equilibria, respectively.

**Figure 2:**
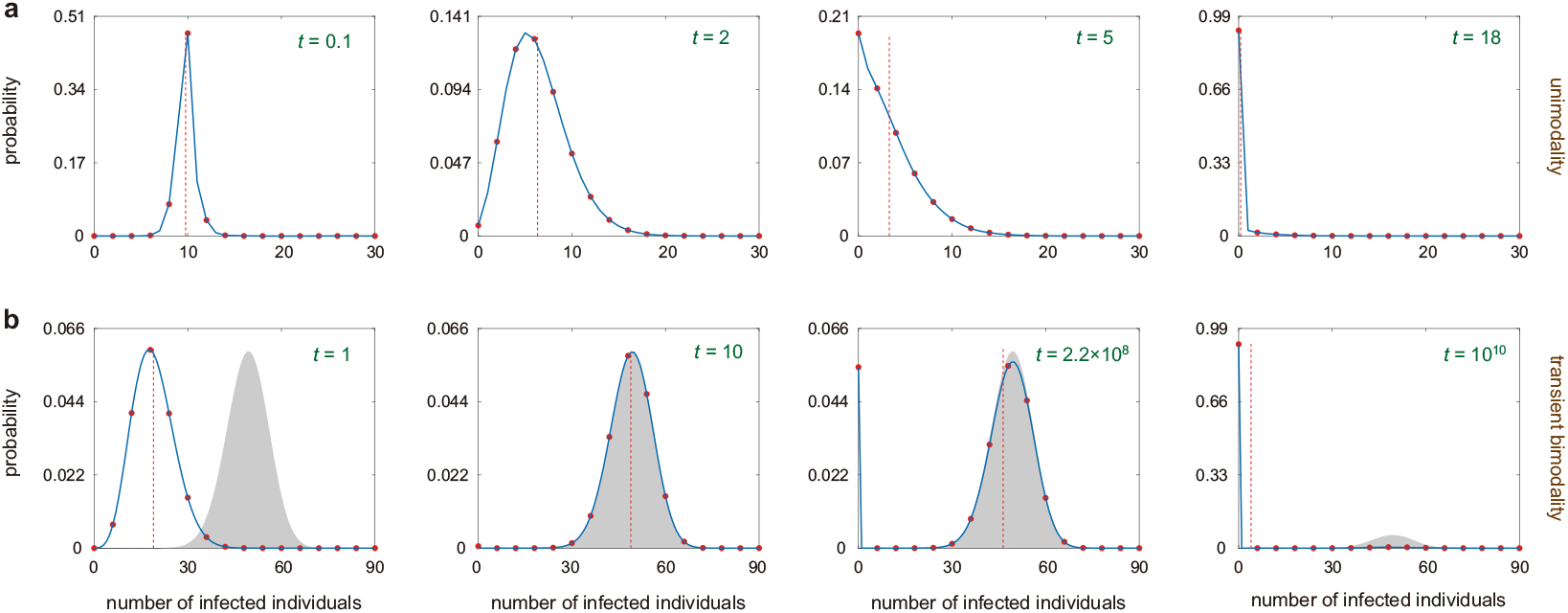
Two distinct patterns of time evolution for the stochastic SIS model. **(a)** When *R*_0_ is small, the distribution of infected individuals is unimodal at small times. The blue curve shows the exact time-dependent distribution given by Eq. (10), the red circles show the numerical one obtained from stochastic simulations, and the red dashed vertical line shows the mean number of infected individuals. The model parameters are chosen as *α* = 0.3, *β* = 0.06, *γ* = 0.6, *R*_0_ = 0.7, and *N* = 100. **(b)** When *R*_0_ is large, the distribution of infected individuals becomes bimodal with both a zero and a non-zero peak at intermediate times and then reverts back to being unimodal at large times. Prior to disease extinction, the distribution can be approximately stationary. The grey region shows the quasi-stationary distribution of infected individuals computed using Eq. (19). The model parameters are chosen as *α* = 0.8, *β* = 0.4, *γ* = 0.6, *R*_0_ = 2.67, and *N* = 100. In (a),(b), the initial condition is chosen as *N*_*I*_ (0) = 0.1*N*.

In practice, the distribution of the number of infected individuals can be measured through parallel observations across multiple regions. Specifically, suppose that the transmission range of the disease can be divided into several sub-regions of similar size and with comparable epidemiological characteristics, such as different cities or communities implementing identical containment strategies. The numbers of infected individuals in these sub-regions can be approximately regarded as statistically independent and identically distributed samples, and thus empirical distributions of infected individuals at multiple discrete time points can be statistically obtained. The exact time-dependent distribution derived in this paper can then be used to construct a likelihood function, allowing all model parameters to be inferred via the maximum likelihood method [16, 45]. In this way, our analytical results enable the integration of observed data with the theoretical model to achieve accurate parameter inference.

### 3.2 Convergence to the extinction state

We have seen that the disease must eventually go extinct for the stochastic SIS model since the state space is finite with 0 ∈ *S* being the unique absorbing state. Here we will validate this fact using the exact time-dependent solution. By the Perron-Frobenius theorem [46], when the system is ergodic, the generator matrix *Q* must have zero as its eigenvalue with multiplicity one and all other eigenvalues must have negative real parts, i.e.

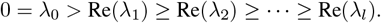

In fact, we can prove a stronger result. Let 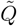 be the matrix obtained from *Q* by removing the first row and the first column. Since the first row of *Q* is the zero vector, all eigenvalues of 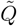 are exactly all nonzero eigenvalues of *Q*. It has been proved in [47] that if all non-absorbing states form a communicating class (this is automatically satisfied for the stochastic SIS model), then the eigenvalue of 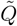 with the maximal real part, i.e. the eigenvalue λ_1_, must be real and has multiplicity one. This indicates that all the eigenvalues of *Q* can be arranged so that

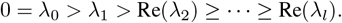

Therefore, the only term in Eq. (10) independent of time *t* is the first term and all other terms tend to zero exponentially fast as *t* → ∞. Since the system is ergodic, the stationary solution of the master equation is independent of the choice of the initial state. As a result, we can take *n*_0_ = *N*, allowing us to focus on to the case of *n*_0_ ≥ *n*, which is now

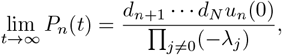

where we have used the fact that *v*_*N*_ (0) = 1. Since the term 1*/*∏ _*j* ≠0_(−λ_*j*_) contributes the same to each term, we can treat it as a normalizing factor. We can also do the same with *d*_*n*+1_ · · · *d*_*N*_, which contributes the same as (*d*_1_ · · · *d*_*n*_)^−1^ up to normalization. Hence the stationary solution has the following simplified expression:

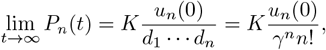

where *K* is a normalization constant. By induction, it is easy to see that *u*_*n*_(0) = *δ*_0_(*n*), where *δ*_0_(*n*) is a Kronecker delta which takes the value of 1 when *n* = 0 and takes the value of 0 otherwise. Since *u*_*n*_(0) = 0 for any *n* ≥ 1, we finally obtain

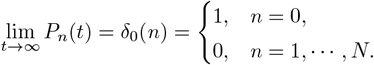

This demonstrates that the stationary distribution of the stochastic SIS model is the point distribution at *n* = 0, corresponding to the extinction state, and the time-dependent distribution converges to this point distribution in the long-term limit.

We emphasize that there is an remarkable difference between the classical and extended SIS models. For the classical SIS model (*β* = 0), the Markovian model shown in Fig. 1(b) reduces to a birth-death process and the generator matrix *Q* given in Eq. (5) reduces to a tridiagonal matrix. In this case, it is well known that the eigenvalues of *Q* are all real numbers [48]. However, for the extended SIS model (*β* ≠ 0), the first two eigenvalues, λ_0_ and λ_1_, are always real, while all other eigenvalues can be complex numbers. To validate this, we compute the first eight eigenvalues of *Q* under different choices of *β* (Table 1). Clearly, all the eigenvalues are real when *β* = 0, as expected. The number of complex eigenvalues increases with the increase of *β*, while the first two eigenvalues are always real.

**Table 1:** The first eight eigenvalues of the generator matrix *Q* for the stochastic SIS model as *β* increases. The model parameters are chosen as *α* = 0.1, *γ* = 1, and *N* = 7.

| eigenvalues | $R_0$ | $\lambda_0$ | $\lambda_1$ | $\lambda_2$ | $\lambda_3$ | $\lambda_4$ | $\lambda_5$ | $\lambda_6$ | $\lambda_7$ |
| --- | --- | --- | --- | --- | --- | --- | --- | --- | --- |
| $\beta = 0$ | 0.1 | 0 | -0.92 | -1.90 | -2.93 | -4.02 | -5.15 | -6.33 | -7.56 |
| $\beta = 0.5$ | 1 | 0 | -0.52 | -1.89 | -3.45 | -5.17 | -5.83 | -6.80+1.78i | -6.80-1.78i |
| $\beta = 12$ | 24 | 0 | -0.002 | -6.61 | -9.07 | -11.36+3.42i | -11.36-3.42i | -15.20+6.53i | -15.20-6.53i |
| $\beta = 24$ | 48 | 0 | -0.0002 | -12.32+1.20i | -12.32-1.20i | -16.60+2.28i | -16.60-2.28i | -25.48+6.64i | -25.48-6.64i |

### 3.3 Dynamical phase diagram and stochastic bifurcation

Recall that the deterministic SIS model has only one equilibrium (disease-free equilibrium) when *R*_0_ ≤ 1 and has two equilibria (disease-free and endemic equilibria) when *R*_0_ *>* 1. A bifurcation occurs at *R*_0_ = 1. Actually, the bifurcation for stochastic epidemic models has also been widely studied using the technique of branching process approximation [23, 24]. It is well known that during the early stage of the disease, the stochastic epidemic model can be approximated by a branching process. This approximation also reveals a bifurcation threshold at *R*_0_ = 1 — the disease undergoes a minor outbreak when *R*_0_ ≤ 1, whereas a major outbreak becomes possible when *R*_0_ *>* 1.

We emphasize that the branching process approximation is only valid during the early stage of the disease. Here we revisit the stochastic bifurcation for the stochastic SIS model by analyzing the time evolution of its probability distribution without relying on any approximations (similar stochastic bifurcation analysis has been performed in stochastic gene networks [49] as well as in stochastic enzyme kinetics [50]). In general, an equilibrium of the deterministic model corresponds to a mode of the probability distribution of the stochastic model. Intuitively, if the deterministic model has only one equilibrium, then the stochastic model should have unimodal distributions at all time points; if the deterministic has two equilibria, then the stochastic model may have bimodal distributions at certain time points — the two modes of the bimodal distribution correspond to the two equilibria of the deterministic model.

We next investigate the shape of the time-dependent distribution for the stochastic SIS model. Following previous studies [23, 24], we assume that initially there is only one infected individual, i.e. *N*_*I*_ (0) = 1. According to simulations, we find that the stochastic SIS model can only produce a unimodal distribution with only one peak or a bimodal distribution with both a zero and a nonzero peak. A distribution with more than two peaks is not observed. We stress that the time-dependent distribution of the number of infected individuals, *P*_*n*_(*t*), must be unimodal when *t* is very small or when *t* is very large. The distribution is unimodal at small times because the initial distribution of the system is the point distribution at *n* = 1, which is unimodal. The distribution is unimodal at large times because the stationary distribution is the point distribution at *n* = 0 (the extinction state), which is also unimodal.

To further understand the shape of the time-dependent distribution, we classify the dynamic behavior of the stochastic SIS model into two different phases: (i) the distribution remains unimodal at all times (Fig. 2(a)); (ii) the distribution is unimodal at small and large times and is bimodal at intermediate times (Fig. 2(b)). Roughly speaking, the two modes of the bimodal distribution correspond to the disease-free and endemic equilibria of the deterministic model, respectively. To distinguish between them, we refer to (i) as unimodality and (ii) as transient bimodality. Note that unimodality corresponds to a minor outbreak of the disease and bimodality corresponds to a major outbreak.

To determine the regions of the two phases in the parameter space, we illustrate the *β/α* − *R*_0_ phase diagram of the stochastic SIS model in Fig. 3 under three different choices of the total population size *N*. Here *β/α* is the ratio of the simultaneous infection rate to the normal infection rate and *R*_0_ is the basic reproduction number. Intriguingly, we find that the system undergoes a stochastic bifurcation from unimodality to transient bimodality as *R*_0_ increases; the bifurcation threshold of *R*_0_ always exceeds 1 and increases as *β/α* increases. Note that this is different from the predictions of the branching process approximation, where *R*_0_ = 1 is the bifurcation point. From Fig. 3(a)-(c), we can see that the bifurcation threshold of *R*_0_ decreases and approaches 1 with the increase of *N*. The indicates that the stochastic bifurcation reduces to the deterministic bifurcation when the population size is very large. Our results also suggest that deterministic models of epidemic dynamics may underestimate the threshold for disease outbreak when the population size is relatively small.

**Figure 3:**
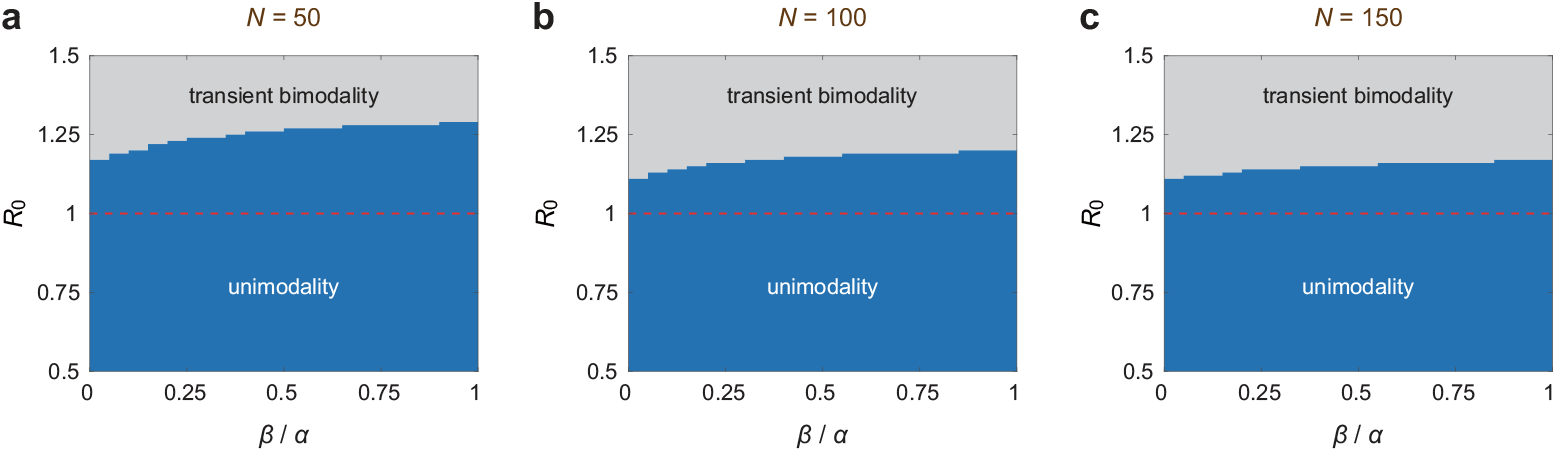
Dynamical phase diagrams for the stochastic SIS model in the *β/α* − *R*_0_ plane as the total population size size *N* increases. The system can produce two different patterns of time evolution: (i) unimodality (blue region) where the distribution is unimodal at small times, and (ii) transient bimodality (grey region) where the distribution becomes bimodal at intermediate times and then reverts back to being unimodal at long times. **(a)** Case of *N* = 50. **(b)** Case of *N* = 100. **(c)** Case of *N* = 150. In (a)-(c), the model parameters are chosen as *γ* = 0.6 and the initial condition is chosen as *N*_*I*_ (0) = 1.

## 4 Quasi-stationary distribution

### 4.1 Exact solution

Recall that when *R*_0_ *>* 1, the deterministic SIS model has an unstable disease-free equilibrium and a positive endemic equilibrium, which is globally asymptotically stable. According to Kurtz’s theorem, when the total population size *N* is large, the trajectory of the stochastic SIS model fluctuates around the endemic equilibrium *x*^∗^ for an extended period before eventually leading to extinction over a much longer timescale (Fig. 1(d)). Prior to disease extinction, the probability distribution of the stochastic SIS model can be approximately stationary. This approximately stationary distribution is widely known as the quasi-stationary (or metastable) distribution [31, 47].

Mathematically, the quasi-stationary distribution is defined as the stationary distribution of the stochastic SIS model conditioned on non-extinction. Note that, given that the disease has not gone extinct at time *t*, the probability distribution of the number of infected individuals at time *t* is given by

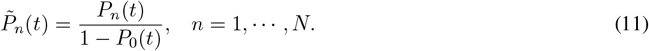

Taking derivatives on both sides yields

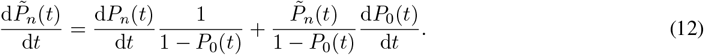

For convenience, we write the generator matrix as *Q* = (*q*_*mn*_)_0≤*m,n*≤*N*_, where *q*_*mn*_, *m* ≠ *n* is the transition rate from state *m* to state *n* and *q*_*mm*_ = − ∑_*m*≠*n*_ *q*_*mn*_. It then follows from Eq. (4) that

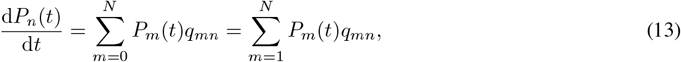

where we have used the fact that *q*_0*n*_ = 0 since the first row of *Q* is the zero vector. Moreover, applying Eq. (4) yields

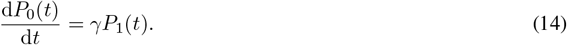

Inserting Eqs. (13) and (14) into Eq. (12), we obtain

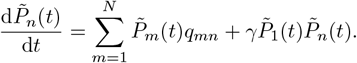

Therefore, conditioned on non-extinction, the evolution of the distribution 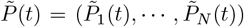 (*t*)) of infected individuals is governed by

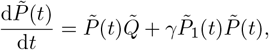

where 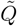 is the matrix obtained from *Q* by deleting the first row and the first column. It has been proved [47] that this system has a unique stationary solution

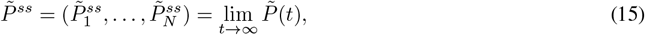

which is referred to as the quasi-stationary distribution of the stochastic SIS model. Clearly, the quasi-stationary distribution 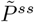 must satisfy

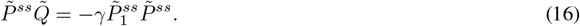

Note that Eq. (16) is difficult to solve since it is nonlinear. We now use the exact time-dependent distribution obtained previously to compute the quasi-stationary distribution. Note that the convergence in Eq. (15) is independent of the choice of the initial state. Hence we can take *n*_0_ = *N*, allowing us to focus on to the case of *n*_0_ ≥ *n*. It then follows from Eq. (10) that

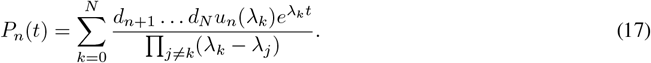

In particular, taking *n* = 0 in the above equation yields

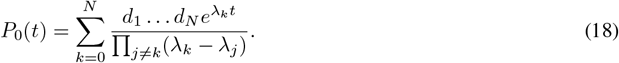

Inserting Eqs. (17) and (18) into Eq. (11), we obtain the exact time-dependent distribution of infected individuals conditioned on non-extinction:

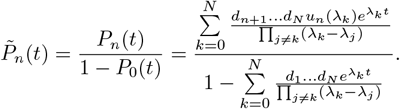

It then follows from Eq. (15) that

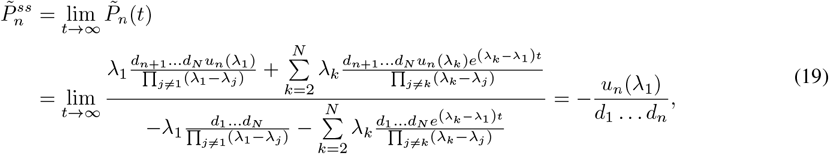

where we have used the fact that λ_1_ *>* Re(λ_2_) … ≥ Re(λ_*N*_). This gives the explicit expression of the quasi-stationary distribution of infected individuals. In particular, it follows from Eq. (19) that

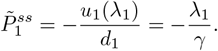

This equation, together with Eq. (16), yields

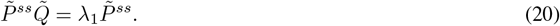

This clearly demonstrates that the quasi-stationary distribution is exactly the left eigenvector of 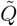 associated with the eigenvalue λ_1_. From Fig. 2(b), we can see that when *R*_0_ is large, the time-dependent distribution of infected individuals initially approaches the quasi-stationary distribution at a relatively short timescale, then becomes bimodal, and eventually leads to extinction at a much longer timescale.

### 4.2 Stochastic bifurcation

Fig. 4(a) shows the quasi-stationary distribution for the stochastic SIS model computed using Eq. (19). Interestingly, we find that the quasi-stationary distribution may have two different shapes. When *β* is small, the quasi-stationary distribution is always unimodal (red solid curve). However, when *β* is large, the distribution may be bimodal with a peak at *n* = 1 and the other peak at *n >* 1 (blue solid curve).

**Figure 4:**
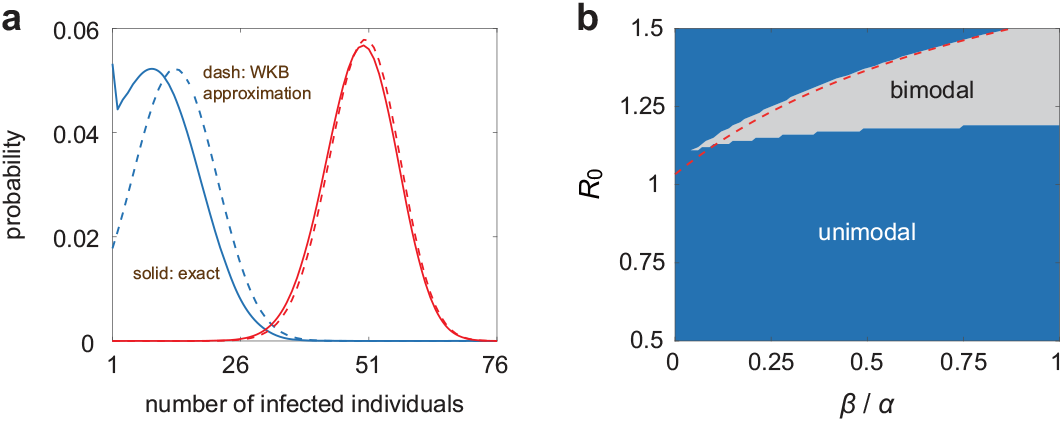
Quasi-stationary distribution of infected individuals for the stochastic SIS model. **(a)** Comparison between the exact quasi-stationary distribution (blue and red solid curves) given by Eq. (19) and the WKB approximation (blue and red dashed curves) computed using Eq. (21). The model parameters are chosen as *α* = 0.12, *β* = 0.33, *γ* = 0.6, *R*_0_ = 1.3, and *N* = 100 for the blue curves, and are chosen as *α* = 1.1, *β* = 0.11, *γ* = 0.6, *R*_0_ = 2.2, and *N* = 100 for the red curves. **(b)** Phase diagrams for the quasi-stationary distribution in the *β/α − R*_0_ plane. The system can produce two different shapes of quasi-stationary distributions: unimodal (blue region) and bimodal (grey region). The red dashed line shows the graph of the curve *R*_0_ = (1 + 2*β/α*)*/*(1 + *β/α*). Theoretically, it can be proved that the bimodal region is (approximately) below this red dashed line. The model parameters are chosen as *γ* = 0.6 and *N* = 100.

To determine the regions for the two distinct shapes of quasi-stationary distributions in the parameter space, we illustrate the *β/α* − *R*_0_ phase diagram in Fig. 4(b). It is clear that the quasi-stationary distribution is always unimodal for the classical SIS model (*β* = 0). When *β* is relatively large, we find that the extended SIS model may display a bimodal distribution when *R*_0_ is neither too large nor too small. Recall that when *R*_0_ is very large, the system produces transient bimodality (Fig. 3(b)) and hence the distribution of infected individuals must be bimodal at at least one time point. However, the quasi-stationary distribution is always unimodal when *R*_0_ is very large. This is a crucial difference between the quasi-stationary distribution and the time-dependent distribution.

To explain why the quasi-stationary distribution is always unimodal when *R*_0_ is very large, note that if the quasi-stationary distribution is bimodal, then it must has a peak at *n* = 1, implying that 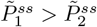. From Eq. (19), the condition of 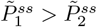 is equivalent to

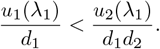

From Eq. (9), this can be rewritten as

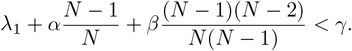

When *R*_0_ is very large, the first nonzero eigenvalue λ_1_ can be ignored, i.e. λ_1_ ≈ 0 (Table 1), and hence the above condition reduces to

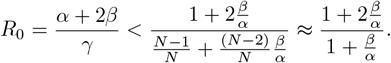

This indicates that a bimodal quasi-stationary only takes place when *R*_0_ *<* (1 + 2*β/α*)*/*(1 + *β/α*) (shown by the region under the red dashed curve in Fig. 4(b)) and hence fails to be observed when *R*_0_ is very large. In summary, we find that the quasi-stationary distribution tends to be bimodal when *β* is relatively large and when *R*_0_ is neither too large nor too small. In addition, comparing Figs. 3(b) and 4(b), we find that the bimodal region of quasi-stationary distribution is much smaller than the region of transient bimodality. This is intuitive because if the quasi-stationary distribution is bimodal, then the distribution of infected individuals must be bimodal at at least one time point and hence the system exhibits transient bimodality.

### 4.3 WKB approximation

Thus far, we have computed the quasi-stationary distribution of the stochastic SIS model in closed form. However, the explicit expression given in Eq. (19) is complicated. Next we apply the WKB theory to derive an approximate analytical solution in the limit of large population size. For simplicity, here we only focus on the case of *R*_0_ *>* 1. In this case, the deterministic model has a disease-free equilibrium which is unstable and a positive endemic equilibrium which is globally asymptotic stable. For convenience, let *x* = *n/N* denote the density of infected individuals and we employ the WKB ansatz

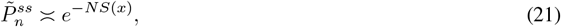

where *S*(*x*) is called the quasi-potential of the stochastic SIS model. The rationality of this ansatz can be rigorously established by using Freidlin-Wentzell-type large deviation theory [51, 52]. In Supplementary Sec. 1, we prove that the quasi-potential is given by

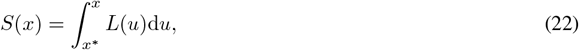

where *x*^∗^ *>* 0 is the endemic equilibrium of the deterministic system and

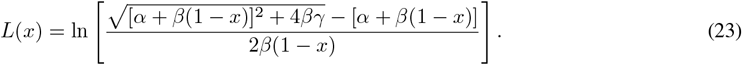

Note that Eqs. (21) and (22) give an approximate analytical expression for the quasi-stationary distribution.

From Eq. (23), it is easy to check that *L*^*′*^(*x*) *>* 0 for any *x* ≥ 0 and *L*(*x*^∗^) = 0. This shows that *L*(*x*) is strictly increasing, *L*(*x*) ≤ 0 for any *x* ≤ *x*^∗^, and *L*(*x*) *>* 0 for any *x > x*^∗^. It then follows from Eq. (21) that

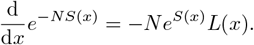

Hence *e*^−*NS*(*x*)^ is monotonically increasing when *x* ≤ *x*^∗^ and is monotonically decreasing when *x* ≥ *x*^∗^. This clearly shows that the WKB approximation of the quasi-stationary distribution must be unimodal.

To evaluate the performance of the WKB approximation, we compare it with the exact quasi-stationary distribution obtained from Eq. (19) in Fig. 4(a). We find that the WKB approximation is in excellent agreement with the exact solution when the quasi-stationary distribution is unimodal (shown by the red solid and dashed lines). However, it may deviate significantly from the exact solution when the quasi-stationary distribution is bimodal (shown by the blue solid and dashed lines). We have proved that the WKB approximation is always unimodal and hence fails to capture the bimodal characteristics of the quasi-stationary distribution.

## 5 Extinction time distribution

### 5.1 Relaxation rate for the deterministic model

We have seen that the disease must eventually go extinct for the stochastic SIS model. Next we investigate the extinction rate and extinction time for the stochastic dynamics. Before doing this, we first focus on the relaxation rate for the deterministic model.

Recall that the deterministic SIS model given in Eq. (2) has an unique global asymptotic stable equilibrium *x*_*e*_, which is zero when *R*_0_ ≤ 1 and is positive when *R*_0_ *>* 1. In the long-term limit, the deterministic system converges to this equilibrium at exponential speed: there exists a constant *C >* 0 such that

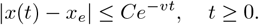

Here the optimal (maximal) parameter *v* characterizes the exponential converge rate to the equilibrium *x*_*e*_, which is also called the relaxation rate in the physics literature [53]. We next compute the relaxation rate *v* analytically. To this end, we make the ansatz

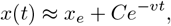

which is a good approximation when *t* is sufficiently large. Inserting it into Eq. (2) yields

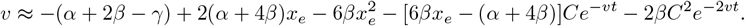

Taking *t* → ∞ in the above equation, we obtain

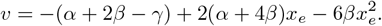

Note that *x*_*e*_ = 0 is the disease-free equilibrium when *R*_0_ ≤ 1 and *x*_*e*_ = *x*^∗^ is the endemic equilibrium when *R*_0_ *>* 1; hence we obtain the following explicit expression of the relation rate:

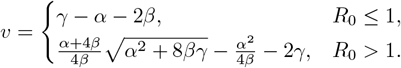

We next investigate the connection between the relaxation rate *v* for the deterministic model and the eigenvalues of the generator matrix *Q* for the stochastic model. Recall that *Q* must have the zero eigenvalue *λ*_0_ = 0. Fig. 5(a) illustrates the relaxation rate *v*, the absolute value of the first nonzero eigenvalue of *Q* (we have seen that the first nonzero eigenvalue must be real and negative), i.e. |*λ*_1_|, and the absolute value of the real part of the second nonzero eigenvalue of *Q*, i.e. |Re(*λ*_2_)|, as a function of the basic reproduction number *R*_0_. It is clear that the relaxation rate *v* for the deterministic model decreases with *R*_0_ when *R*_0_ ≤ 1 and increases with *R*_0_ when *R*_0_ *>* 1. Interestingly, we find that the relaxation rate *v* is very close to |*λ*_1_| when *R*_0_ is relatively small, whereas it is very closed to |Re(*λ*_2_)| when *R*_0_ is relatively large. This provides deep insights into the first and second nonzero eigenvalues of the generator matrix for the stochastic SIS model. Roughly speaking, for the stochastic dynamics, the first nonzero eigenvalue *λ*_1_ characterizes the rate of disease extinction (since |*λ*_1_| ≈ *v* = *γ*(1 − *R*_0_) when *R*_0_ *<* 1), i.e. the convergence rate to the extinction state, while the second nonzero eigenvalue *λ*_2_ characterizes the rate of disease outbreak (since |Re(*λ*_2_)| ≈ *v* when *R*_0_ *>* 1), i.e. the convergence rate to the quasi-stationary state.

**Figure 5:**
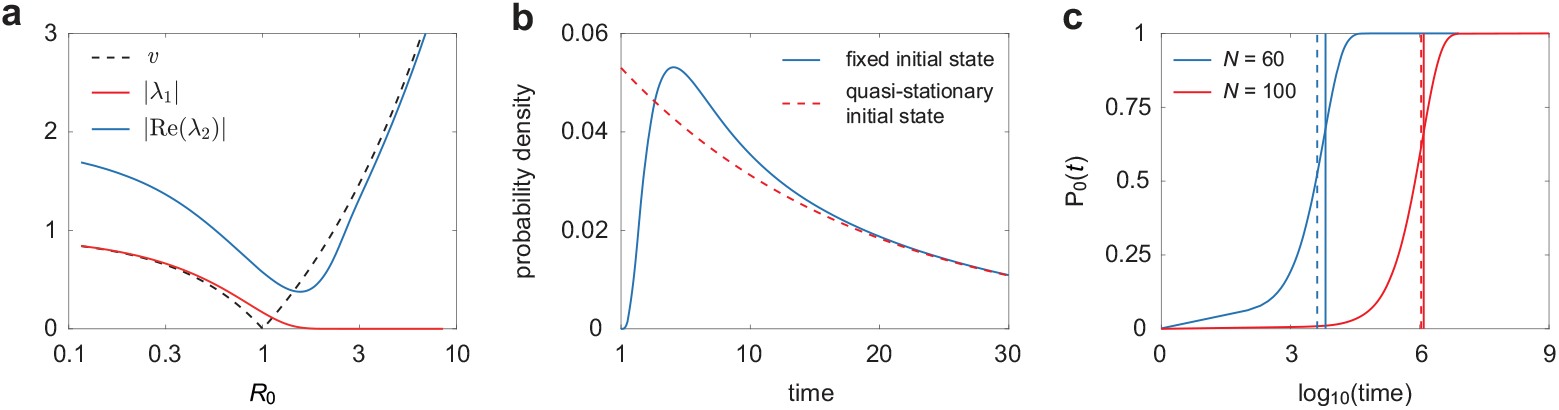
Extinction rate and extinction time for the stochastic SIS model. **(a)** Comparison of the relaxation rate *v* to the global asymptotically stable equilibrium for the deterministic model, the absolute value of the first nonzero eigenvalue |*λ*_1_| for the stochastic model, and the absolute value of the real part of the second nonzero eigenvalue |Re(*λ*_2_) | for the stochastic model as *R*_0_ varies. The model parameters are chosen as *α* = 0.09, *γ* = 0.95, and *N* = 100. **(b)** Comparison of the extinction time distributions when the system starts from a fixed initial state (blue curve) and when the system starts from the quasi-stationary distribution (red dashed curve). The model parameters are chosen as *α* = 0.5, *β* = 0.1, *γ* = 0.6, *R*_0_ = 1.17, and *N* = 60. **(c)** Comparison the exact and approximate mean extinction times. The blue and red curves show the time evolution of *P*_0_(*t*) under two different choices of *N*. The solid vertical lines show the exact mean extinction time computed using Eq. (26) and the dashed vertical lines show the WKB approximation computed using Eq. (27). The model parameters are chosen as *α* = 0.8, *β* = 0.2, *γ* = 0.6, and *R*_0_ = 2. In (b),(c), the initial condition is chosen to be *N*_*I*_ (0) = 0.1*N*.

As a summary, we emphasize that the stationary distribution (extinction state) of the stochastic SIS model corresponds to the eigenvector associated with the zero eigenvalue *λ*_0_ and the convergence rate to the extinction state is characterized by the first nonzero eigenvalue *λ*_1_. Similarly, the quasi-stationary distribution of the stochastic SIS model corresponds to the eigenvector associated with the first nonzero eigenvalue *λ*_1_ and the convergence rate to the quasi-stationary state is characterized by the second nonzero eigenvalue *λ*_2_.

### 5.2 Exact extinction time distribution

We next exmaine the extinction dynamics of the disease. Let *T* = inf{*t* ≥ 0 : *N*_*I*_ (*t*) = 0} denote the time to extinction for the stochastic SIS model. We next compute the distribution of the extinction time. Since *n* = 0 is the absorbing state, it is easy to see that the events *T* ≤ *t* and *N*_*I*_ (*t*) = 0 are equivalent. The probability of *N*_*I*_ (*t*) = 0 has been obtained analytically in Eq. (18). Hence it is easy to compute the distribution of the extinction time *T*. Suppose that the initial number of infected individuals is *n*_0_ ≥ 1. It follows from Eq. (10) that

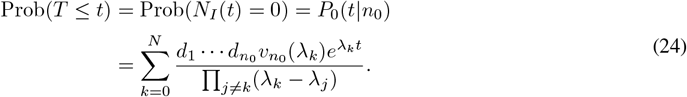

Differentiating this equation with respect to *t*, we obtain the explicit expression of the probability density *f* (*t*) of the extinction time *T* :

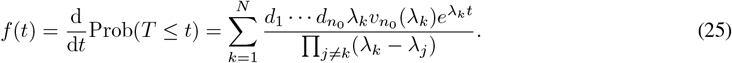

The gives the exact extinction time distribution when the system starts from a fixed initial state.

In some cases, it is more important to study the extinction time distribution starting from the quasi-stationary state rather than from a fixed initial state. We next compute the extinction time distribution when the system starts from the quasi-stationary distribution 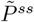. Since 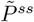 is stationary solution of Eq. (12), we have

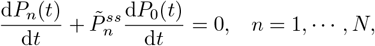

where we have used the fact that 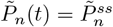 for any *t* ≥ 0 and *n* ≥ 1 when the system starts from the quasi-stationary distribution. Moreover, it follows from Eq. (14) that 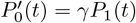. This indicates that

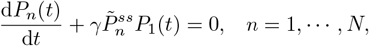

Taking *n* = 1 in the above equation, we obtain

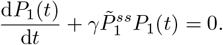

Solving this equation yields 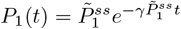. Since 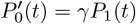, we finally obtain the probability density *f* (*t*) of the extinction time *T* when the system starts from the quasi-stationary distribution:

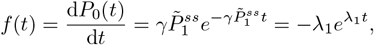

where we have used the fact that 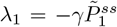. This demonstrates that the extinction time follows an exponential distribution with rate −*λ*_1_ when the system starts from the quasi-stationary distribution. This is consistent with the results obtained in [47].

Fig. 5(b) compares the extinction time distributions when the system starts from the quasi-stationary distribution and when it starts from a fixed initial state. In the former case, the extinction time distribution is exponential and thus monotonically decreasing (shown by the red dashed curve). In the latter case, the extinction time distribution may be non-exponential and bell-shaped, increasing initially and then decreasing (shown by the blue curve).

### 5.3 Exact mean extinction time and the WKB approximation

We next compute the mean extinction time for the stochastic SIS model. We first consider the case where the initial number of infected individuals is *n*_0_ ≥ 1. From Eq. (25), the mean extinction time is given by

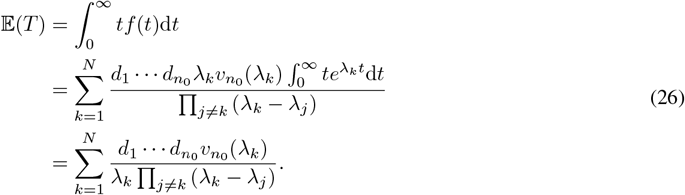

This gives the explicit expression of the mean extinction time when the system starts from a fixed initial state. When the system starts from the quasi-stationary distribution, we have proved that the extinction time follows an exponential distribution with parameter −*λ*_1_. Hence the mean extinction time is *E* (*T*) = −1*/λ*_1_. This again shows that the first nonzero eigenvalue of *Q* characterizes the convergence speed to the extinction state.

Thus far, we have obtained the mean extinction time for the stochastic SIS model. However, the explicit expression given in Eq. (26) is complicated and it is not clear how the population size affects the mean extinction time. Next we use the WKB theory to derive an approximate analytical solution of the mean extinction time when the total population size is large. For simplicity, here we only focus on the case of *R*_0_ *>* 1. According to the Freidlin-Wentzell-type large deviation theory [51, 52], the leading-order asymptotic behaviour of the mean extinction time is given by

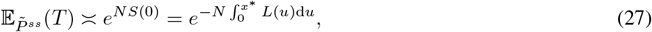

where *S*(0) = *S*(0) − *S*(*x*^∗^) represents the energy barrier between the disease-free equilibrium, i.e. the extinction state, and the endemic equilibrium *x*^∗^, and the function *L*(*u*) is given by Eq. (23). Clearly, the mean extinction time increases exponentially with the total population size *N*.

To validate the analytical results, we focus on the evolution of *P*_0_(*t*) when the system starts from a fixed initial state. Fig. 5(c) illustrates the time evolution of *P*_0_(*t*) obtained from Eq. (24), the exact mean extinction time, and the approximate mean extinction time predicted using the WKB theory under different population sizes. It can be seen that when *N* is large, *P*_0_(*t*) (shown by the colored curve) is close to zero at small times and is close one at large times. In particular, there is a sharp critical transition of *P*_0_(*t*) from zero to one at intermediate times. We emphasize that the increase of *P*_0_(*t*) is due to disease extinction, and hence the critical transition must occur around the mean extinction time. We find that the approximate mean extinction time predicted by the WKB theory (shown by the dashed vertical line) is very closed to the exact mean extinction time (shown by the solid vertical line). This suggests that the WKB approximation indeed provides an excellent prediction of the mean extinction time.

## 6 First-passage time distribution

In addition to the extinction time, public health authorities are particularly concerned with the time at which the number of infected individuals first reaches or exceeds a specified threshold — an indicator known as the first-passage time. This indicator provides a quantitative basis for constructing early warning systems and supports the design of graded response strategies. Moreover, it plays a critical role in guiding both the timing and intensity of intervention measures, thereby helping to delay the epidemic peak and optimize the allocation of resources [54, 55]. Specifically, let *T*_*M*_ = inf*{t* : *N*_*I*_ (*t*) ≥ *M }* denote the time at which the number of infected individuals first reaches or exceeds a given threshold *M* ≥ 1. We next compute the distribution of the first-passage time *T*_*M*_ for the stochastic SIS model. Suppose that the initial number of infected individuals is *n*_0_ ∈ [1, *M*) and let

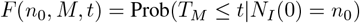

be the cumulative distribution function of *T*_*M*_. Note that within the small time interval [0, Δ*t*], the system can only make three types of transitions (Fig. 1(b)): a susceptible individual is infected with probability 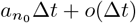, two susceptible individuals are simultaneously infected with probability 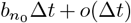, and an infected individual recovers with probability 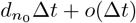. On the other hand, it is clear that the probability of having more than one transitions during [0, Δ*t*] is *o*(Δ*t*), and the probability of having no transitions is 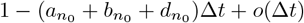. Hence it follows that

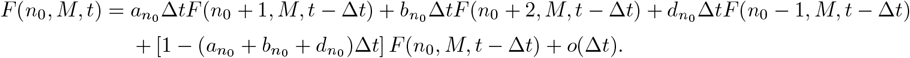

Subtracting *F* (*n*_0_, *M, t* − Δ*t*) on both sides of the above equation and letting Δ*t* → 0, we find that *F* (*n*_0_, *M, t*) is governed by the following Kolmogorov backward equation:

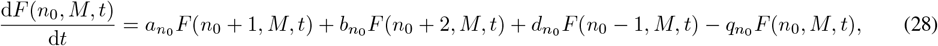

where the boundary conditions are given by *F* (0, *M, t*) = 0 and *F* (*n*_0_, *M, t*) = 1 for any *n*_0_ ≥ *M*. To proceed, let

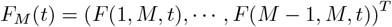

be a column vector whose components are *F* (*n*_0_, *M, t*) for all *n*_0_ ∈ [1, *M*). Then Eq. (28) can be written in matrix form as

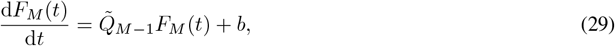

where *b* = (0, 0, …, *b*_*M*−2_, *a*_*M*−1_ + *b*_*M*−1_)^*T*^ and

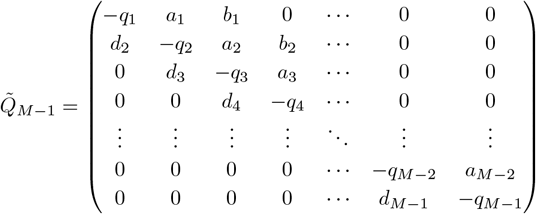

is the matrix obtained from 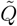 by retaining its first *M* − 1 rows and *M* − 1 columns. Since *F*_*M*_ (0) = 0 is the zero vector, it follows from Eq. (29) that

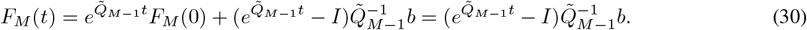

Similarly to the derivation of the exact time-dependent distribution, we compute 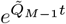 and 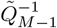 analytically using matrix-valued Cauchy’s integral formula and Cauchy’s residue theorem. Then *F*_*M*_ (*t*) can be computed in closed form as (see Supplementary Sec. 2 for the proof):

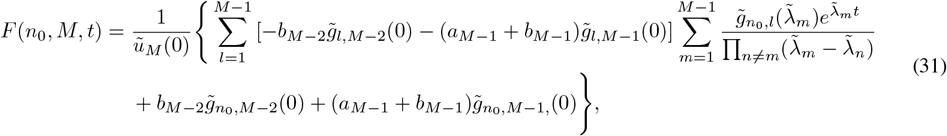

where 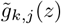 are polynomials defined as

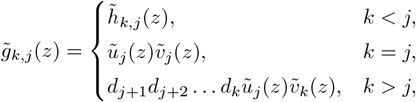

and 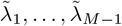 are all the eigenvalues of 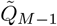 (for simplicity, we assume that these eigenvalues are non-degenerate and hence 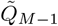 has *M* − 1 distinct eigenvalues). Here 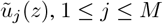, 1 ≤ *j* ≤ *M*, are polynomials defined recursively by

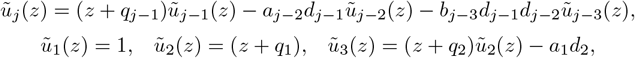

with 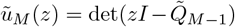 being the characteristic polynomial of 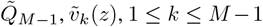, are polynomials defined recursively by

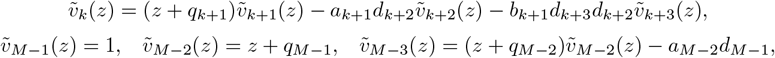

and 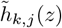 are functions defined as

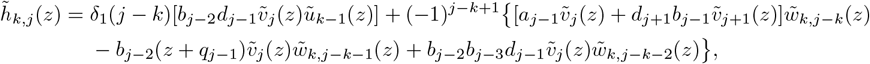

Where 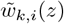, 1 ≤ *i* ≤ *j* − *k*, are another sequence of polynomials defined recursively by

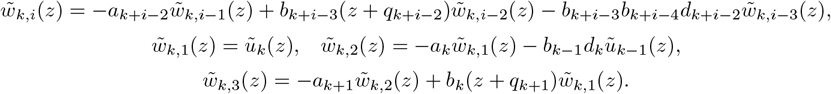

Differentiating Eq. (31) with respect to *t*, we finally obtain the probability density of the first-passage time *T*_*M*_ :

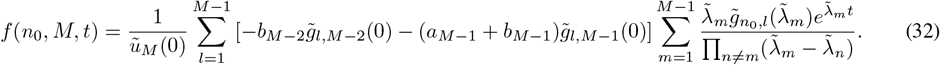

As expected, the analytical solution coincides perfectly with the numerical one obtained using the stochastic simulation algorithm (Fig. 6(a)). Moreover, we emphasize that for any given threshold *M > n*_0_, there is a positive probability that the disease will have gone extinct before the number of infected individuals ever reaches *M*. Consequently, the first-passage time *T*_*M*_ can take an infinite value with positive probability, implying that its expected value must diverge to infinity, i.e. *E* [*T*_*M*_] = ∞.

**Figure 6:**
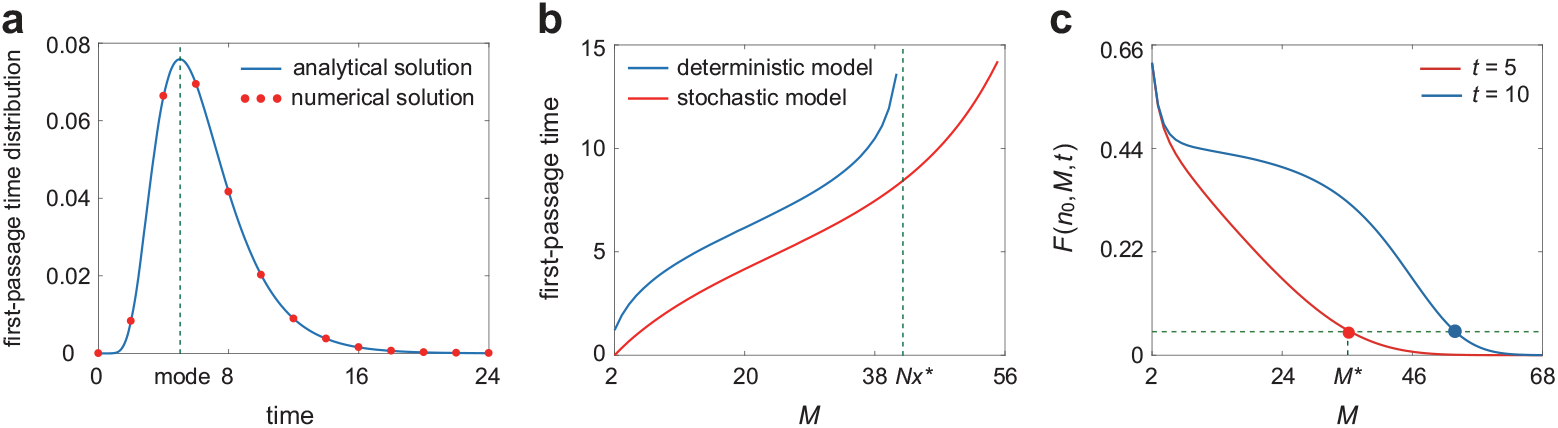
First-passage time distribution for the stochastic SIS model. **(a)** Comparison of the analytical first-passage time distribution given in Eq. (32) (blue curve) and the numerical one (red dots) obtained from stochastic simulations. The green dashed vertical line shows the mode (peak value) of the distribution. The model parameters are chosen as *N* = 100, *α* = 0.8, *β* = 0.2, *γ* = 0.6, and *R*_0_ = 2. The initial condition is chosen as *N*_*I*_ (0) = 1 and the critical threshold is chosen as *M* = 30. **(b)** Comparison of the first-passage time for the deterministic model (blue curve) and the mode of the first-passage time distribution for the stochastic model (red curve) as the threshold *M* varies. The green dashed vertical line shows the endemic equilibrium *Nx*^∗^ for the deterministic model. **(c)** The probability that the number of infections has already reached the threshold *M* before time *t*, i.e. *F* (*n*_0_, *M, t*), as a function of *M*. Given the significance level of *α* = 0.05, the critical threshold *M* ^∗^ satisfying *F* (*n*_0_, *M* ^∗^, *t*) = *α* gives an estimate of the emergency bed capacity at time *t* (with 95% confidence). In (b),(c), the model parameters and initial condition are chosen to be the same as in (a).

Fig. 6(b) compares the first-passage time for the deterministic model with the mode of the first-passage time distribution for the stochastic model (we use the mode here rather than the mean since the expected value of *T*_*M*_ diverges to infinity), both as functions of the threshold *M*. Clearly, the deterministic model predicts a longer first-passage time than the stochastic model. In particular, the discrepancy becomes pronounced when the threshold *M* approaches the endemic equilibrium *Nx*^∗^. This occurs because the deterministic system can never actually reach the endemic equilibrium, leading to an infinite first-passage time when *M* ≥ *Nx*^∗^, whereas the stochastic system still yields a finite value. In the context of infectious disease control, the first-passage time at which the number of infections reaches a critical threshold is often regarded as a warning signal for intervention measures. As shown in Fig. 6(b), the deterministic model may severely overestimate the first-passage time, potentially delaying critical response actions and thereby increasing the scale and risk of disease outbreaks. By contrast, the stochastic model provides a more timely and reliable warning signal for interventions.

Next we demonstrate how analyzing the first-passage time distribution can inform the allocation of emergency beds. Fig. 6(c) illustrates *F* (*n*_0_, *M, t*) as a function of *M* for two different values of *t*, where *F* (*n*_0_, *M, t*) denotes the probability that the number of infections has already reached the threshold *M* by time *t*. For any given significance level *α >* 0, one can determine the critical threshold *M* ^∗^ satisfying *F* (*n*_0_, *M* ^∗^, *t*) = *α* (Fig. 6(c)). For example, if we choose *α* = 0.05, then with 95% probability the number of infections will not have reached *M* ^∗^ by time *t*. This provides practical guidance for short-term healthcare resource planning: by setting the emergency bed capacity to *M* ^∗^, hospitals can meet disease control demand at time *t* with 95% confidence. Thus analysis of the first-passage time for the stochastic SIS model offers a quantitative basis for early warning and control.

## 7 Conclusions and discussion

In this work, we provide a systematic study of the stochastic dynamics of an extended SIS epidemic model in densely populated environments using a Markov jump process framework. Unlike the classical SIS model, which only includes the infection of a susceptible individual and the recovery of an infected individual, the extended SIS model allows for simultaneous transmission of the disease to two susceptible individuals due to crowded conditions. It is well known that the deterministic SIS model has a positive endemic equilibrium when the basic reproduction number is greater than one. However, the stochastic SIS model will eventually go extinct with probability one, since the state space is finite and the extinction state is an absorbing state of the system.

Using complex analysis techniques including matrix-valued Cauchy’s integral formula and Cauchy’s residue theorem, we analytically solve the master equation for the stochastic SIS model and derive closed-form solutions for the time-dependent distribution of the number of infected individuals as well as the distribution of the first-passage time at which the number of infected individuals reaches a certain threshold. Prior to disease extinction, the stochastic SIS model approaches an approximately stationary state corresponding to the endemic equilibrium of the deterministic model. This analytical time-dependent distribution is then used to compute the quasi-stationary distribution of infected individuals and the extinction time distribution of the disease. Moreover, the quasi-potential of the stochastic SIS model is derived using the large deviation theory, which is then used to construct WKB approximations of the quasi-stationary distribution and the mean extinction time in the limit of large population size.

Within the Markov jump process framework, the generator matrix for the classical SIS model has only real eigenvalues. In contrast, the extended SIS model has a zero eigenvalue and a real first nonzero eigenvalue, but all other eigenvalues may be complex due to the simultaneous infection of two susceptible individuals. The extinction state is shown to correspond to the eigenvector associated with the zero eigenvalue, while the quasi-stationary state corresponds to the eigenvector associated with the first nonzero eigenvalue. We also investigate the extinction and outbreak rates of the infectious disease. Interestingly, we find that the first nonzero eigenvalue of the generator matrix characterizes the extinction rate of the epidemic, while the second nonzero eigenvalue characterizes the outbreak rate.

We also study the stochastic bifurcation for the stochastic SIS model by analyzing the time evolution of the distribution of infected individuals, and find that the system may exhibit two distinct dynamic patterns: (i) unimodality, where the distribution remains unimodal at all times; and (ii) transient bimodality, where the distribution is unimodal at early and late times but becomes bimodal at intermediate times. The bifurcation threshold of the basic reproduction number for the stochastic model is shown to be larger than that for its deterministic counterpart. Similarly, we examine the shape of the quasi-stationary distribution and find that it may be unimodal or bimodal. The parameter region for bimodality in the quasi-stationary distribution is shown to be much narrower than that for transient bimodality. Finally, we demonstrate that analyzing the first-passage time distribution provides a quantitative basis for early warning of intervention measures and rational planning of medical resources, such as hospital beds and drug inventories, thereby improving the efficiency of resource allocation and emergency response capabilities.

Our study also has some limitations. First, we only consider the case where at most two susceptible individuals can be infected simultaneously. However, in densely populated settings or during highly contagious phases of an outbreak, more than two individuals may become infected simultaneously. Second, to ensure analytical tractability, we have not considered the birth of susceptible individuals and the death of susceptible and infected ones [56]. Such extensions would lead to a more complex Markov process, for which analytical treatment may be much more challenging. We aim to explore these more complex scenarios in future work to gain deeper insights into infectious disease dynamics and improve epidemic control strategies. We also anticipate that our analytical results can be used to perform accurate parameter inference for stochastic epidemic dynamics based on empirical probability distributions obtained from multi-region parallel observations at multiple discrete time points.

## Data Availability

All data produced in the present work are contained in the manuscript

## Acknowledgements

G. Liu acknowledges support from NSFC grants Nos.12371494, 12231012, and 11971279, as well as Shanxi Provincial Key Research and Development Project No.202202020101010. C. Jia acknowledges support from NSFC grants No. 12271020.

