## Supplementary Material for "Exact results for the stochastic SIS epidemic model in densely populated environments"

#### Contents

**1 WKB approximation of the quasi-stationary distribution** 2

**2 Derivation of the exact first-passage time distribution** 3

### 1 WKB approximation of the quasi-stationary distribution

Here we apply the WKB theory to derive an approximate analytical solution of the quasi-stationary distribution in the limit of large population size. For simplicity, here we only focus on the case of $R_0 > 1$. In this case, the deterministic model has a disease-free equilibrium which is unstable and a positive endemic equilibrium which is globally asymptotic stable. When the total population size $N$ is large, the trajectory of the stochastic SIS model fluctuates around the endemic equilibrium for an extended period before eventually leading to extinction over a much longer timescale (Fig. 1(d) in the main text). This implies that the first nonzero eigenvalue $\lambda_1$ of the generator matrix $Q$ must be sufficient small when $N \gg 1$ since it characterizes the convergence rate to the extinction state. This is also evident from Fig. 5(a) in the main text, where $\lambda_1$ is indeed very small and thus can be ignored when $R_0 > 1$. In this case, the right-hand size in Eq. (20) in the main text can be ignored and hence we obtain $\tilde{P}^{ss}\tilde{Q} = 0$, i.e.

$$a_{n-1}\tilde{P}^{ss}_{n-1} + b_{n-2}\tilde{P}^{ss}_{n-2} + d_{n+1}\tilde{P}^{ss}_{n+1} - q_n\tilde{P}^{ss}_n = 0, \quad n = 1, \cdots, N. \tag{1}$$

For convenience, let $x = n/N$ denote the density of infected individuals. When $N \gg 1$, it is clear that the transition rates $a_n$, $b_n$, and $d_n$ for the stochastic SIS model have the following approximations:

$$\begin{aligned} a_n/N &= \alpha x(1-x) := a(x), \\ b_n/N &\approx \beta x(1-x)^2 := b(x), \\ d_n/N &= \gamma x := d(x). \end{aligned}$$

We then employ the WKB ansatz

$$\tilde{P}^{ss}_n \asymp e^{-NS(x)},$$

where $S(x)$ is called the quasi-potential of the stochastic SIS model. The rationality of this ansatz can be rigorously established by using Freidlin-Wentzell-type large deviation theory [1–3]. Inserting this ansatz into Eq. (1) yields

$$\begin{aligned} &a(x-1/N)e^{-NS(x-1/N)} + b(x-2/N)e^{-NS(x-2/N)} \\ &+ d(x+1/N)e^{-NS(x+1/N)} - [a(x)+b(x)+d(x)]e^{-NS(x)} = 0. \end{aligned}$$

Taylor expanding the above equation with respect to $1/N$, and collecting the leading order terms yields

$$a(x)\left(e^{S'(x)} - 1\right) + b(x)\left(e^{2S'(x)} - 1\right) + d(x)\left(e^{-S'(x)} - 1\right) = 0.$$

To proceed, we introduce the Hamiltonian

$$H(x,p) = a(x)\left(e^p - 1\right) + b(x)\left(e^{2p} - 1\right) + d(x)\left(e^{-p} - 1\right).$$

Then it is clear that the quasi-potential $S(x)$ satisfies the following time-independent Hamilton Jacobi equation:

$$H(x, S'(x)) = 0. \tag{2}$$

We emphasize that the extended SIS model is equivalent to the reaction system given in Eq. (1) in the main text. In fact, the Hamiltonian for a general chemical reaction network was first obtained in [4].

We next solve the Hamilton-Jacobi equation. Note that Eq. (2) can be rewritten in a simpler form as

$$\left(e^{S'(x)} - 1\right)\left[a(x) + b(x)\left(e^{S'(x)} + 1\right) - d(x)e^{-S'(x)}\right] = 0.$$

Hence we have $e^{S'(x)} = 1$ or

$$a(x) + b(x)\left(e^{S'(x)} + 1\right) - d(x)e^{-S'(x)} = 0. \tag{3}$$

Note that $e^{S'(x)} = 1$ implies that $S(x)$ is a constant. This solution is trivial and hence is of no interest to us. The nontrivial solution can be obtained by solving Eq. (3) and is given by

$$S(x) = \int_{x^*}^{x} L(u)\mathrm{d}u,$$

where $x^*$ is the endemic equilibrium of the deterministic system and

$$\begin{aligned} L(x) =& \ln\left[\frac{[(a(x)+b(x))^2 + 4b(x)d(x)]^{\frac{1}{2}} - (a(x)+b(x))}{2b(x)}\right] \\ =& \ln\left[\frac{\sqrt{(\alpha+\beta(1-x))^2 + 4\beta\gamma} - (\alpha+\beta(1-x))}{2\beta(1-x)}\right]. \end{aligned}$$

According to the Freidlin-Wentzell-type large deviation theory [1–3], the approximate analytical expression for the quasi-stationary distribution is given by

$$\tilde{P}_n^{ss} \asymp e^{-NS(x)} = e^{-N\int_{x^*}^{x} L(u)\mathrm{d}u}.$$

### 2 Derivation of the exact first-passage time distribution

Let $F(n_0, M, t) = \text{Prob}(T_M \leq t | N_I(0) = n_0)$ denote the cumulative distribution function of the first-passage time $T_M$ given that the initial number of infected individuals is $n_0 \geq 1$, and let

$$F_M(t) = (F(1, M, t), \cdots, F(M-1, M, t))^T$$

be a column vector whose components is composed of $F(n_0, M, t)$ for all $n_0 \in [1, M)$. In the main text, we have shown that $F_M(t)$ has the following expression:

$$F_M(t) = (e^{\tilde{Q}_{M-1}t} - I)\tilde{Q}_{M-1}^{-1}b, \tag{4}$$

where $b = (0, 0, \ldots, b_{M-2}, a_{M-1} + b_{M-1})^T$ and

$$\tilde{Q}_{M-1} = \begin{pmatrix} -q_1 & a_1 & b_1 & 0 & \cdots & 0 & 0 \\ d_2 & -q_2 & a_2 & b_2 & \cdots & 0 & 0 \\ 0 & d_3 & -q_3 & a_3 & \cdots & 0 & 0 \\ 0 & 0 & d_4 & -q_4 & \cdots & 0 & 0 \\ \vdots & \vdots & \vdots & \vdots & \ddots & \vdots & \vdots \\ 0 & 0 & 0 & 0 & \cdots & -q_{M-2} & a_{M-2} \\ 0 & 0 & 0 & 0 & \cdots & d_{M-1} & -q_{M-1} \end{pmatrix}$$

is the matrix obtained from $\tilde{Q}$ by retaining its first $M-1$ rows and $M-1$ columns.

We next compute $F_M(t)$ in closed form. Let $r_{k,j}(t)$ and $\eta_{k,j}$ be the $(k, j)$th element of $e^{\tilde{Q}_{M-1}t}$ and $\tilde{Q}_{M-1}^{-1}$, respectively. It then follows from Eq. (4) that

$$\begin{aligned} F(n_0, M, t) =& \sum_{l=1}^{M-1} \left[b_{M-2}\eta_{l,M-2} + (a_{M-1} + b_{M-1})\eta_{l,M-1}\right] r_{n_0,l}(t) \\ & - \left[b_{M-2}\eta_{n_0,M-2} + (a_{M-1} + b_{M-1})\eta_{n_0,M-1}\right]. \end{aligned} \tag{5}$$

We still use matrix-valued Cauchy's integral formula and Cauchy's residue theorem from Sec. 3.1 in the main text to calculate $r_{k,j}(t)$. By Cauchy's integral formula for matrices, for any continuous function $f$, we have

$$f(\tilde{Q}_{M-1}) = \frac{1}{2\pi i}\oint_C f(z)(zI - \tilde{Q}_{M-1})^{-1}\mathrm{d}z,$$

where $C$ is an arbitrary simple closed curve in the complex plane that contains all the eigenvalues of $\tilde{Q}_{M-1}$ in its interior. If we take $f(z) = e^{zt}$, then we obtain

$$e^{\tilde{Q}_{M-1}t} = \frac{1}{2\pi i}\oint_C (zI - \tilde{Q}_{M-1})^{-1}e^{zt}\mathrm{d}z.$$

Thus, we yield

$$r_{k,j}(t) = \frac{1}{2\pi i}\oint_C (zI - \tilde{Q}_{M-1})^{-1}_{k,j}e^{zt}\mathrm{d}z, \tag{6}$$

where $(zI - \tilde{Q}_{M-1})^{-1}_{k,j}$ denotes the $(k, j)$th element of the matrix $(zI - \tilde{Q}_{M-1})^{-1}$. By Cramer's rule of calculating the inverse matrix, we can prove that

$$(zI - \tilde{Q}_{M-1})^{-1}_{k,j} = \frac{1}{\tilde{u}_M(z)}\begin{cases}\tilde{h}_{k,j}(z), & k < j,\\ \tilde{u}_j(z)\tilde{v}_j(z), & k = j,\\ d_{j+1}\dots d_k\tilde{u}_j(z)\tilde{v}_k(z), & k > j,\end{cases} \tag{7}$$

where $\tilde{u}_j(z)$, $1 \le j \le M$, are polynomials defined recursively by

$$\tilde{u}_j(z) = (z + q_{j-1})\tilde{u}_{j-1}(z) - a_{j-2}d_{j-1}\tilde{u}_{j-2}(z) - b_{j-3}d_{j-1}d_{j-2}\tilde{u}_{j-3}(z),$$
$$\tilde{u}_1(z) = 1, \quad \tilde{u}_2(z) = (z + q_1), \quad \tilde{u}_3(z) = (z + q_2)\tilde{u}_2(z) - a_1d_2,$$

with $\tilde{u}_M(z) = \det(zI - \tilde{Q}_{M-1})$ being the characteristic polynomial of $\tilde{Q}_{M-1}$, $\tilde{v}_k(z)$, $1 \le k \le M-1$, are polynomials defined recursively by

$$\tilde{v}_k(z) = (z + q_{k+1})\tilde{v}_{k+1}(z) - a_{k+1}d_{k+2}\tilde{v}_{k+2}(z) - b_{k+1}d_{k+3}d_{k+2}\tilde{v}_{k+3}(z),$$
$$\tilde{v}_{M-1}(z) = 1, \quad \tilde{v}_{M-2}(z) = z + q_{M-1}, \quad \tilde{v}_{M-3}(z) = (z + q_{M-2})\tilde{v}_{M-2}(z) - a_{M-2}d_{M-1},$$

and $\tilde{h}_{k,j}(z)$ are functions defined as

$$\tilde{h}_{k,j}(z) = \delta_1(j-k)[b_{j-2}d_{j-1}\tilde{v}_j(z)\tilde{u}_{k-1}(z)] + (-1)^{j-k+1}\big\{[a_{j-1}\tilde{v}_j(z) + d_{j+1}b_{j-1}\tilde{v}_{j+1}(z)]\tilde{w}_{k,j-k}(z)$$
$$- b_{j-2}(z + q_{j-1})\tilde{v}_j(z)\tilde{w}_{k,j-k-1}(z) + b_{j-2}b_{j-3}d_{j-1}\tilde{v}_j(z)\tilde{w}_{k,j-k-2}(z)\big\}.$$

Here $\tilde{w}_{k,i}(z)$, $1 \le i \le j-k$, are another sequence of polynomials defined recursively by

$$\tilde{w}_{k,i}(z) = -a_{k+i-2}\tilde{w}_{k,i-1}(z) + b_{k+i-3}(z + q_{k+i-2})\tilde{w}_{k,i-2}(z) - b_{k+i-3}b_{k+i-4}d_{k+i-2}\tilde{w}_{k,i-3}(z),$$
$$\tilde{w}_{k,1}(z) = \tilde{u}_k(z), \quad \tilde{w}_{k,2}(z) = -a_k\tilde{w}_{k,1}(z) - b_{k-1}d_k\tilde{u}_{k-1}(z),$$
$$\tilde{w}_{k,3}(z) = -a_{k+1}\tilde{w}_{k,2}(z) + b_k(z + q_{k+1})\tilde{w}_{k,1}(z).$$

Combining Eqs. (6) and (7), we obtain

$$r_{k,j}(t) = \frac{1}{2\pi i}\oint_C \frac{\tilde{g}_{k,j}(z)}{\tilde{u}_M(z)}e^{zt}\mathrm{d}z,$$

where $\tilde{g}_{k,j}(z)$ are polynomials defined as

$$\tilde{g}_{k,j}(z) = \begin{cases} \tilde{h}_{k,j}(z), & k < j, \\ \tilde{u}_j(z)\tilde{v}_j(z), & k = j, \\ d_{j+1}d_{j+2}\dots d_k\tilde{u}_j(z)\tilde{v}_k(z), & k > j. \end{cases}$$

Since $\tilde{u}_M(z)$ is the characteristic polynomial of $\tilde{Q}_{M-1}$, we have

$$\tilde{u}_M(z) = (z-\tilde{\lambda}_1)^{\tilde{r}_1}\dots(z-\tilde{\lambda}_s)^{\tilde{r}_s},$$

where $\tilde{\lambda}_1,\dots,\tilde{\lambda}_s$ are all pairwise distinct eigenvalues of $\tilde{Q}_{M-1}$ with $\tilde{r}_1,\dots,\tilde{r}_s$ being their multiplicities, respectively.

To proceed, we apply Cauchy's residue theorem

$$\oint_C f(z)\mathrm{d}z = 2\pi i\sum_k \mathrm{Res}(f;\varepsilon_k),$$

where $\varepsilon_k$ are all isolated singularities of $f$ inside the simple closed curve $C$. In our current case, all the isolated singularities are the eigenvalues $\tilde{\lambda}_m$ and thus

$$\oint_C \frac{\tilde{g}_{k,j}(z)}{\tilde{u}_M(z)}e^{zt}\mathrm{d}z = 2\pi i\sum_{m=1}^{s}\frac{1}{(\tilde{r}_m-1)!}\frac{\mathrm{d}^{\tilde{r}_m-1}}{\mathrm{d}z^{\tilde{r}_m-1}}\left.\frac{\tilde{g}_{k,j}(z)e^{zt}}{\prod_{n\neq m}(z-\tilde{\lambda}_n)^{\tilde{r}_n}}\right|_{z=\tilde{\lambda}_m}.$$

Therefore, once we have known all the eigenvalues of $\tilde{Q}_{M-1}$, we obtain

$$r_{k,j}(t) = \sum_{m=1}^{s}\frac{1}{(\tilde{r}_m-1)!}\frac{\mathrm{d}^{\tilde{r}_m-1}}{\mathrm{d}z^{\tilde{r}_m-1}}\left.\frac{\tilde{g}_{k,j}(z)e^{zt}}{\prod_{n\neq m}(z-\tilde{\lambda}_n)^{\tilde{r}_n}}\right|_{z=\tilde{\lambda}_m}.$$

In most cases, the eigenvalues of $\tilde{Q}_{M-1}$ are mutually different. In this case, we have $s = M-1$ and $\tilde{r}_1 = \dots = \tilde{r}_s = 1$, and thus the time-dependent distribution can be simplified as

$$r_{k,j}(t) = \sum_{m=1}^{M-1}\frac{\tilde{g}_{k,j}(\tilde{\lambda}_m)e^{\tilde{\lambda}_m t}}{\prod_{n\neq m}(\tilde{\lambda}_m-\tilde{\lambda}_n)}, \tag{8}$$

where $\tilde{\lambda}_1,\dots,\tilde{\lambda}_{M-1}$ are all the eigenvalues of $\tilde{Q}_{M-1}$, i.e. all zeros of the characteristic polynomial $\det(zI-\tilde{Q}_{M-1})$.

Next we calculate $\eta_{k,j}$. Since $\tilde{Q}_{M-1}^{-1} = -(zI-\tilde{Q}_{M-1})^{-1}|_{z=0}$, we obtain

$$\eta_{k,j} = -\tilde{g}_{k,j}(0)/\tilde{u}_M(0).$$

This shows that

$$\eta_{l,M-2} = -\tilde{g}_{l,M-2}(0)/\tilde{u}_M(0), \quad \eta_{l,M-1} = -\tilde{g}_{l,M-1}(0)/\tilde{u}_M(0). \tag{9}$$

Inserting Eqs. (8) and (9) into Eq. (5), we obtain the analytical expression of $F(n_0,M,t)$, which is given by

$$F(n_0,M,t) = \frac{1}{\tilde{u}_M(0)}\Bigg\{\sum_{l=1}^{M-1}\left[-b_{M-2}\tilde{g}_{l,M-2}(0)-(a_{M-1}+b_{M-1})\tilde{g}_{l,M-1}(0)\right]\sum_{m=1}^{M-1}\frac{\tilde{g}_{n_0,l}(\tilde{\lambda}_m)e^{\tilde{\lambda}_m t}}{\prod_{n\neq m}(\tilde{\lambda}_m-\tilde{\lambda}_n)}$$
$$+ b_{M-2}\tilde{g}_{n_0,M-2}(0)+(a_{M-1}+b_{M-1})\tilde{g}_{n_0,M-1}(0)\Bigg\}.$$
